# Voices from the frontline: findings from a thematic analysis of a rapid online global survey of maternal and newborn health professionals facing the COVID-19 pandemic

**DOI:** 10.1101/2020.05.08.20093393

**Authors:** Aline Semaan, Constance Audet, Elise Huysmans, Bosede B Afolabi, Bouchra Assarag, Aduragbemi Banke-Thomas, Hannah Blencowe, Severine Caluwaerts, Oona M R Campbell, Francesca L Cavallaro, Leonardo Chavane, Louise Tina Day, Alexandre Delamou, Therese Delvaux, Wendy Graham, Giorgia Gon, Peter Kascak, Mitsuaki Matsui, Sarah G Moxon, Annettee Nakimuli, Andrea B Pembe, Emma Radovich, Thomas van den Akker, Lenka Benova

## Abstract

**Objective:** To prospectively document experiences of frontline maternal and newborn healthcare providers during the COVID-19 pandemic.

**Design:** Cross-sectional study via an online survey disseminated through professional networks and social media in 12 languages. We analysed responses using descriptive statistics and qualitative thematic analysis disaggregating by low- and middle-income countries (LMICs) and high-income countries (HICs).

**Setting:** 81 countries, between March 24 and April 10, 2020.

**Participants:** 714 maternal and newborn healthcare providers.

**Main outcome measures:** Preparedness for and response to COVID-19, experiences of health workers providing care to women and newborns, and adaptations to 17 outpatient and inpatient care processes during the pandemic.

**Results:** Only one third of respondents received training on COVID-19 from their health facility and nearly all searched for information themselves. Half of respondents in LMICs received updated guidelines for care provision compared with 82% in HICs. Overall, only 47% of participants in LMICs, and 69% in HICs felt mostly or completely knowledgeable in how to care for COVID-19 maternity patients. Facility-level responses to COVID-19 (signage, screening, testing, and isolation rooms) were more common in HICs than LMICs. Globally, 90% of respondents reported somewhat or substantially higher levels of stress. There was a widespread perception of reduced use of routine maternity care services, and of modification in care processes, some of which were not evidence-based.

**Conclusions:** Substantial knowledge gaps exist in guidance on management of maternity cases with or without COVID-19. Formal information sharing channels for providers must be established and mental health support provided. Surveys of maternity care providers can help track the situation, capture innovations, and support rapid development of effective responses.

**Key Messages:**

What is already known
- In addition to lack of healthcare worker protection, staffing shortages, heightened risk of nosocomial transmission and decreased healthcare use described in previous infectious disease outbreaks, maternal and newborn care during the COVID-19 pandemic has also been affected by large-scale lockdowns/curfews.
- The two studies assessing the indirect effects of COVID-19 on maternal and child health have used models to estimate mortality impacts.
- Experiences of frontline health professionals providing maternal and newborn care during the COVID-19 pandemic have not been empirically documented to date.

What this study adds
- Respondents in high-income countries more commonly reported available/updated guidelines, access to COVID-19 testing, and dedicated isolation rooms for confirmed/suspected COVID-19 maternity patients.
- Levels of stress increased among health professionals globally, including due to changed working hours, difficulties in reaching health facilities, and staff shortages.
- Healthcare providers were worried about the impact of rapidly changing care practices on health outcomes: reduced access to antenatal care, fewer outpatient visits, shorter length-of-stay in facilities after birth, banning birth companions, separating newborns from COVID-19 positive mothers, and postponing routine immunisations.
- COVID-19 illustrates the susceptibility of maternity care services to emergencies, including by reversing hard-won gains in healthcare utilisation and use of evidence-based practices. These rapid findings can inform countries of the main issues emerging and help develop effective responses.

## Introduction

Coronavirus disease (COVID-19) is a respiratory tract infection caused by the severe acute respiratory syndrome coronavirus 2 (SARS-CoV-2), which was first recognized in December 2019 in Wuhan, China.^1^ COVID-19 is a highly infectious disease with two main routes of transmission: directly via close contacts with an infected person and indirectly via contact with contaminated surfaces. While evidence gathering continues, concerns are emerging regarding a possible vertical transmission (antenatally or intrapartum).^2,3^ The effect of COVID-19 infection during the 1^st^ and 2^nd^ trimester of pregnancy remains to be clarified, highlighting the need for a good surveillance system to register adverse outcomes arising from infection in early pregnancy. At present, the virus has not been detected in breast milk of mothers with confirmed (or suspected) COVID-19 infection, so transmission via breastfeeding is then considered unlikely. Direct transmission from mother to child may occur via close contact, but breastfeeding continues to be encouraged with appropriate hygiene measures, including wearing face masks.^4,5^ As of May 4^th^ 2020, 3.3 million cases of COVID-19 have been confirmed and more than 230,000 deaths reported globally.^6^ Based on data collected by the Chinese Centre for Disease Control and Prevention, an infection with COVID-19 can cause a range of illness severity: from mild to moderate (>80%), to severe (14%) and critical (5%). The overall case fatality rate is estimated at 2.3%.^7^ Among the risk factors associated with an increased case fatality rate are older age, male sex, and comorbidities, particularly cardiovascular diseases and diabetes.^8^

Studies are currently ongoing to evaluate whether pregnant women have an increased susceptibility to infection with COVID-19 and if they present a greater risk of severe illness or mortality. However, the available limited evidence suggests that pregnant women have risks of infection comparable to the general population.^9^ The disease severity in pregnant women does not appear significantly higher than in non-pregnant women.^10,11^ A meta-analysis conducted by Di Mascio et al. of 41 pregnant women hospitalised in a context for a COVID-19 infection showed an increased risk of preterm birth, preeclampsia, and caesarean section.^12^ Symptoms in newborns suspected or confirmed with COVID-19 seem to be mild, with good outcomes.^13^ though one study reported a higher risk of perinatal death.^12^ Amoroux et al. draw attention to the possible delay in the development of visible hypoxemic lesions in newborns of COVID-19 positive mothers, and advise their close follow-up after birth.^3^ However, the limited size of samples in these studies calls for caution and more data need to be collected to draw definitive conclusions.^14^ Considering the increased risk of infection with other respiratory viruses such as influenza, and the increased mortality linked with H1N1, it is important for pregnant women to be protected from illnesses.^12^ However, for pregnant women, irrespective of the risks, symptoms, and severity, the main recommendations to avoid infection remain the same as for the general public.^15,16^ Some countries, such as the United Kingdom, issued stricter measures for pregnant women, categorising them as part of a vulnerable group applying the precautionary principles and recommending self-isolation.^9^

The effects of COVID-19 are likely to go beyond the direct provision of care to women and newborns with suspected or confirmed COVID-19 infection. Previous infectious disease outbreaks severely reduced the capacity of health systems to provide critical reproductive, maternal, and newborn healthcare, with negative impacts on their health outcomes.^16–18^ Studies of recent outbreaks of Ebola virus disease (EBV), SARS and MERS, highlighted several challenges in countries’ preparedness to face outbreaks, amplified by a weak existing systems. This includes lack of protection and safety for healthcare workers leading to disruption in staffing, heightened risk of nosocomial transmission, and elevated stress levels among service providers.^19,20^ A qualitative study by Qian Liu et al. also shows that healthcare providers in China were stressed during the ongoing outbreak because of an added workload and fear of contracting and transmitting the infection.^21^ Other indirect consequences of previous infectious disease outbreaks include less healthcare utilisation and limited capacity for public health surveillance.^22–24^ These impacts can persist long after the disease outbreak is contained.^25^ However, much of the evidence available about these impacts on maternal and newborn health is either modelled or is from studies using secondary data such as population-based surveys and routine health management information system analysis.^22,26,27^ Additionally, large disruption to health-seeking behaviour and healthcare provision is caused by the unprecedented measures countries implement to contain the pandemic (e.g., lockdowns, curfews, restrictions on public transport).

To date, studies assessing the potential indirect effects of the COVID-19 pandemic on sexual, reproductive, maternal and child health have used modelling approaches. Roberton and colleagues modelled three scenarios projecting a decrease in the coverage of basic life-saving interventions, varying the extent and duration.^28^ They estimated an increase in maternal deaths between 12,190 and 56,700, and 253,500-1,157,000 additional deaths of children under five years of age. Similar conclusions have been drawn by Riley et al. who projected that a modest decline in the use of sexual and reproductive healthcare services in 132 low- and middle-income countries (LMICs) will result, over the course of a year, in 48 million additional women with unmet need for modern contraceptives, 15 million additional unwanted pregnancies, and over 3 million additional unsafe abortions will occur.^29^ It is therefore critical that the precise nature of the direct and indirect impacts of COVID-19, and the adaptations and innovations tested to reduce its impact are captured and described, and that this be done prospectively.^30^

Levels of preparedness and response in maternal and newborn care services differ markedly between institutions and countries. Health personnel, including those defined by the World Health Organization as “competent maternal and newborn health professionals educated, trained and regulated to national and international standards”^31^ are at the frontline of providing care for pregnant women and their newborns during the COVID-19 pandemic. They are expected to be sufficiently competent to: (i) provide and promote evidence-based, human-rights based, quality, socio-culturally sensitive and dignified care to women and newborns; (ii) facilitate physiological processes during labour and delivery to ensure a clean and positive childbirth experience; and (iii) identify and manage or refer women and/or newborns with complications. They are the ones that serve as the interface between the governments working to address the pandemic and pregnant women experiencing the consequences, as such they are uniquely placed to be able to describe the status of care provision during such times.

The objective of this paper is to synthesise the key themes identified in the first round of a global online survey of health professionals working in maternal and newborn health along four dimensions: preparedness for COVID-19, response to COVID-19, personal experience in the workplace, and changes in provision of care and care processes. This online survey is part of a larger study which seeks to 1) understand how health professionals and health facilities prepare and respond to COVID-19 in regard to the care provided to women and their babies during antenatal, intrapartum and postnatal care; and 2) document and analyse the effect of the COVID-19 pandemic on the services available to pregnant, labouring and postpartum women and their newborns, including as a result of increasing pressures on the healthcare system.

## Methods

### Study design

This is a cross-sectional study of health professionals providing maternal and newborn healthcare services. In the future, we plan to collect repeated rounds of the online survey to track the preparedness for, response to and effects of the COVID-19 pandemic over time and follow-up qualitative individual interviews from selected respondents to gain additional insights.

### Population and sampling

The target population for this survey was health professionals directly providing maternal (antenatal, intrapartum and/or postnatal) or newborn care. We included cadres such as midwives, nurses, obstetricians, gynaecologists, neonatologists, paediatricians, anaesthetists, general practitioners, medical officers, clinical officers, community health workers, lactation counsellors, paramedics, health technicians, and others, including health professionals in training. Due to the unavailability of a global sampling frame for this study population, the survey sampling was non-random and not intended to generate generalisable nationally representative results of either health professionals or health facilities. Rather, our intention was to collect and synthesise the voices and experiences of maternal and newborn health professionals from a range of countries, contexts, services and facility types at the early stage of the COVID-19 pandemic. An invitation to complete the survey was distributed using personal networks of the multi-country research team members, maternal/newborn platforms, and social media (e.g. Facebook, Twitter, WhatsApp). Respondents were encouraged to share the survey with other colleagues in an attempt to snowball the sample population. Respondents provided informed consent online by checking a box affirming that they understood the consent form and voluntarily agreed to participate in the survey.

### Questionnaire development

A questionnaire was developed by an international team of collaborators including health professionals, experts in health systems, infectious diseases, infection prevention and control, maternal health epidemiologists, and public health researchers from various global settings. The questionnaire was prepared in English, and translated into 11 languages (French, Arabic, Italian, Portuguese, Spanish, Chinese, Japanese, Russian, German, Swahili, and Dutch), by native speakers with medical training (minimum two translators per language). It was piloted by asking five maternal/newborn health professionals from different global settings to complete the questionnaire and provide feedback. We used this feedback to assess face validity and refine the wording of the questions and the format and wording of response options. We collected data on the respondents’ background (country and region, qualification and work responsibilities, gender, and basic characteristics of the health facility in which the respondents worked, if any). To avoid concerns over confidentiality, we did not collect names of health facilities. The questionnaire included three core modules focusing on preparedness for COVID-19, response to COVID-19, and health workers’ own experience of work during the COVID-19 pandemic. In the fourth, optional module, we asked respondents to elaborate on adaptations to 17 care processes (timing, frequency, modality of contact with patients during various types of outpatient and inpatient care) and to comment on whether they perceived that the uptake of care by the population they serve has changed and, if it had, how. The full questionnaire is provided in Supplementary File 1.

### Data processing and analysis

In this paper, we use responses collected between the day the survey was launched (March 24, 2020) and April 10, 2020. First, we cleaned the 798 responses received by removing duplicate submissions (n=49) and those who did not agree to the consent statement (n=14), and submissions made by those not directly providing maternal or newborn care (such as lecturers and public health officials; n=10). Quantitative analysis involved production of descriptive statistics including frequencies and percentages using Stata/SE version 14, and responses were stratified by country income levels (according to World Bank classification).^32^ We conducted a qualitative thematic analysis of free-text answers to derive common themes related to respondents’ concerns and reported changes in the work environment and care process by country income levels. When possible, we triangulated qualitative and quantitative results to validate emerging themes.

### Missing data

From the 725 remaining responses, we dropped 11 responses with missing answers on more than 90% of the survey questions from analysis. The proportion of missing answers to multiple choice questions ranged between 0.5 and 6.5%, and that to open-ended questions from 16 to 28% of respondents. Missing answers to the “Country” question were recorded based on the “Region” answer for 93 responses; for example, a respondent with a missing response for country but region reported as Maharashtra was coded as from India.

## Results

### Respondents’ characteristics

The analysed sample included a total of 714 healthcare professionals caring for women and newborns, 59% of whom agreed to participate in the optional module of the survey (n=397). Table 1 summarises respondents’ characteristics. Participants were based in 81 countries and more than half (63%) were from high-income countries (HICs). A map showing respondents’ geographic distribution is available in Supplementary File 2. Obstetricians/gynaecologists and midwives constituted the majority of respondents (38% and 35%, respectively), followed by nurse-midwives and nurses. Around one third worked in referral hospitals and 60% were employed in public sector facilities. Most facilities where participants worked provided caesarean sections (81%), accepted referrals from other facilities (71%), and included maternal intensive care units (ICU, 64%) and newborn intensive care units (NICU, 59%). Nearly half of respondents from HICs (49%) reported that their facilities had seen maternity patients with confirmed or suspected COVID-19 infection, compared to 13% of respondents from low-and-middle income countries (LMICs).

**Table 1.** - Survey (n=714^*^) and optional module (n=397) respondent characteristics

|  | Survey (%) | Optional module (%) |
| --- | --- | --- |
| **Country income level (World Bank classification)** |  |  |
| Low and Middle income countries | 263 (37) | 136 (35) |
| High income countries | 444 (63) | 256 (65) |
| **Region** |  |  |
| East Asia & Pacific | 82 (12) | 51 (13) |
| Europe & Central Asia | 249 (35) | 131 (33) |
| Latin America & Caribbean | 43 (6) | 30 (8) |
| Middle East & North Africa | 53 (7) | 29 (7) |
| North America | 87 (12) | 53 (14) |
| South Asia | 83 (12) | 37 (9) |
| Sub-Saharan Africa | 110 (16) | 61 (16) |
| **Cadre** |  |  |
| Midwife | 248 (35) | 135 (34) |
| Nurse-midwife | 83 (12) | 48 (12) |
| Nurse | 22 (3) | 14 (4) |
| Obstetrician/gynaecologist | 269 (38) | 148 (38) |
| Neonatologist | 6 (1) | 3 (1) |
| Paediatrician | 4 (1) | 4 (1) |
| General practitioner | 10 (1) | 5 (1) |
| Medical doctor (no specialization) | 15 (2) | 10 (3) |
| Medical student/intern/resident | 13 (2) | 6 (2) |
| Community health worker/Outreach worker | 12 (2) | 6 (2) |
| Other | 29 (4) | 16 (4) |
| **Position** |  |  |
| Head of facility | 60 (9) | 34 (9) |
| Head of department or ward | 71 (10) | 41 (11) |
| Head of team | 94 (13) | 54 (14) |
| Team member | 346 (50) | 195 (50) |
| Locum or interim member | 22 (3) | 10 (3) |
| Other<sup>a</sup> | 101 (15) | 53 (13) |
| **Type of care provided (multiple responses allowed)** |  |  |
| Outpatient ANC | 438 (61) | 244 (62) |
| Home-based childbirth care | 77 (11) | 47 (12) |
| Outpatient PNC | 316 (44) | 176 (45) |
| Outpatient Breastfeeding support | 217 (30) | 121 (31) |
| Inpatient ANC | 374 (52) | 218 (56) |
| Inpatient childbirth care | 437 (61) | 249 (64) |
| Inpatient PNC | 350 (49) | 193 (50) |
| Surgical care | 213 (30) | 115 (29) |
| Neonatal care (small and sick newborns) | 85 (12) | 47 (12) |
| Home visits | 131 (18) | 78 (20) |
| Community outreach | 105 (15) | 69 (18) |
| Abortion care | 157 (22) | 86 (22) |
| Post-abortion care | 179 (25) | 104 (27) |
| Other | 84 (12) | 42 (11) |
| **Health facility level** |  |  |
| Referral hospital | 250 (36) | 144 (37) |
| District/regional hospital | 154 (22) | 77 (20) |
| Health center | 76 (11) | 46 (12) |
| Polyclinic | 6 (1) | 6 (2) |
| Clinic | 66 (10) | 36 (9) |
| Health post/unit or dispensary | 16 (2) | 9 (2) |
| Other<sup>b</sup> | 116 (17) | 68 (18) |
| **Health facility sector** |  |  |
| Public (national) | 183 (27) | 86 (22) |
| Public (university or teaching) | 138 (20) | 81 (21) |
| Public (district level or below) | 80 (12) | 61 (16) |
| Social security | 7 (1) | 3 (1) |
| Health insurance or HMO | 10 (2) | 7 (2) |
| Private university | 25 (4) | 10 (3) |
| Private for profit | 95 (14) | 57 (15) |
| Non-governmental | 61 (9) | 29 (8) |
| Faith-based or mission | 23 (3) | 15 (4) |
| Other | 56 (8) | 34 (9) |
| **Type of area** |  |  |
| Large city (more than 1 million inhabitants) | 273 (40) | 151 (39) |
| Small city (100,000 to 1 million inhabitants) | 220 (32) | 125 (32) |
| Town (fewer than 100,000 inhabitants) | 106 (16) | 61 (16) |
| Village/Rural area | 64 (9) | 38 (10) |
| Refugee/displaced persons camp | 8 (1) | 2 (1) |
| Other | 9 (1) | 9 (2) |
| **Facility characteristics** |  |  |
| Caesarean-section provision | 535 (81) | 301 (81) |
| Accept referrals from other facilities | 476 (71) | 269 (71) |
| ICU available | 429 (64) | 236 (62) |
| NICU available | 398 (59) | 226 (59) |

\*Differential number of missing values across variables
<sup>a</sup> Mainly self-practicing midwives
<sup>b</sup> Mainly birth centres and private practice
Abbreviations: Antenatal care (ANC); Intensive care unit (ICU); Neonatal intensive care unit (NICU); Postnatal care (PNC)

### Knowledge on the provision of maternal and newborn healthcare and COVID-19

Most respondents (90%) reported that the health facilities where they work provided them with information on preparing for the COVID-19 outbreak (Table 2). This included general guidance about the disease(definition, transmission mode and treatment options), prevention measures (e.g. hand hygiene, disinfecting surfaces and equipment, personal protective equipment [PPE] use, social distancing and isolation), patient screening, case reporting, and updated policies and guidelines. However, only one third of respondents reported receiving hands-on training/drills on the response to COVID-19 (Table 2). Several raised concerns about this lack of access to training activities and perceived this as a necessity that would have made them “*feel better prepared*” to respond to the needs of women and their babies during the outbreak.

**Table 2.**
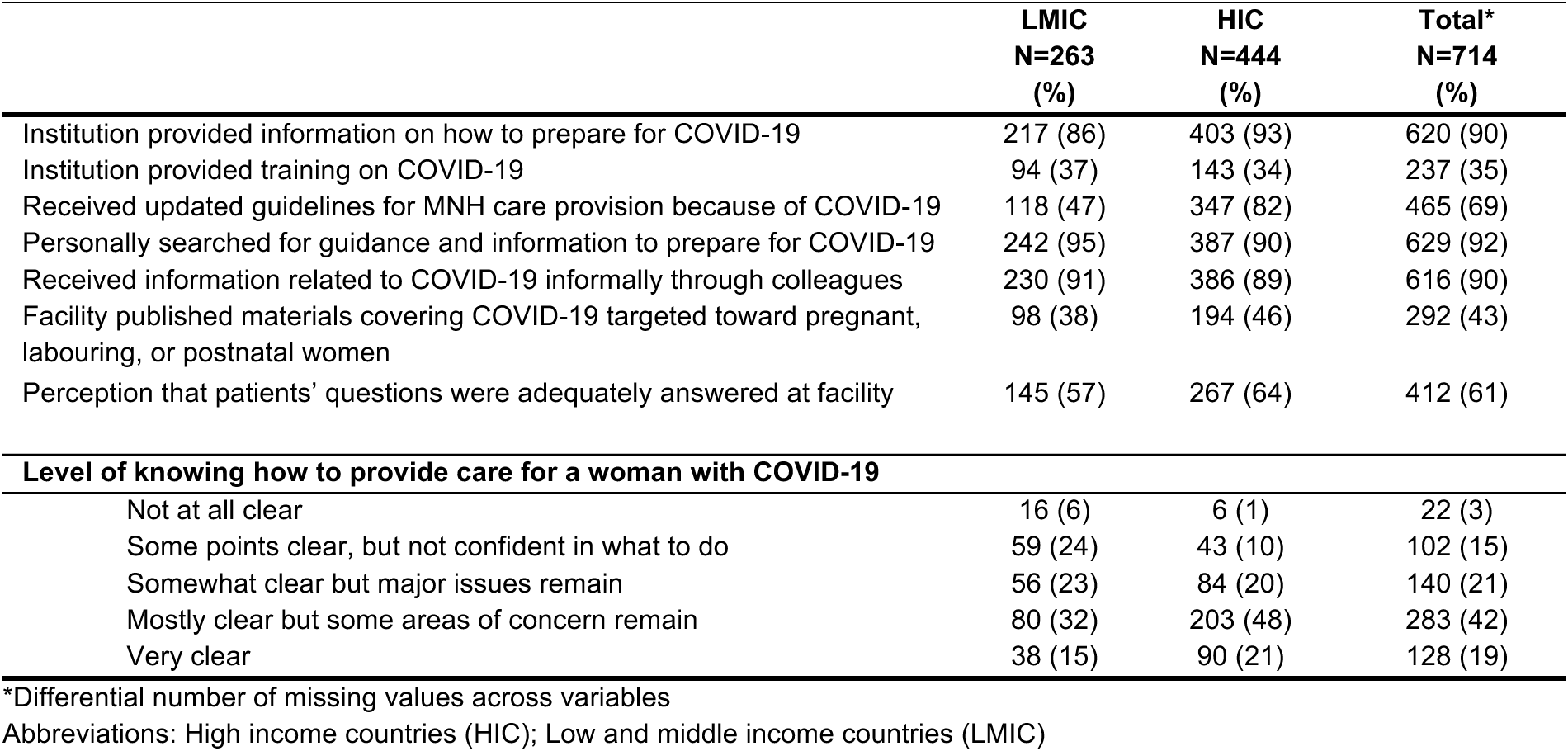
– Preparedness for COVID-19 among maternal and newborn health professionals, by country income category

|  | LMIC<br>N=263<br>(%) | HIC<br>N=444<br>(%) | Total*<br>N=714<br>(%) |
| --- | --- | --- | --- |
| Institution provided information on how to prepare for COVID-19 | 217 (86) | 403 (93) | 620 (90) |
| Institution provided training on COVID-19 | 94 (37) | 143 (34) | 237 (35) |
| Received updated guidelines for MNH care provision because of COVID-19 | 118 (47) | 347 (82) | 465 (69) |
| Personally searched for guidance and information to prepare for COVID-19 | 242 (95) | 387 (90) | 629 (92) |
| Received information related to COVID-19 informally through colleagues | 230 (91) | 386 (89) | 616 (90) |
| Facility published materials covering COVID-19 targeted toward pregnant, labouring, or postnatal women | 98 (38) | 194 (46) | 292 (43) |
| Perception that patients' questions were adequately answered at facility | 145 (57) | 267 (64) | 412 (61) |
| **Level of knowing how to provide care for a woman with COVID-19** |  |  |  |
| Not at all clear | 16 (6) | 6 (1) | 22 (3) |
| Some points clear, but not confident in what to do | 59 (24) | 43 (10) | 102 (15) |
| Somewhat clear but major issues remain | 56 (23) | 84 (20) | 140 (21) |
| Mostly clear but some areas of concern remain | 80 (32) | 203 (48) | 283 (42) |
| Very clear | 38 (15) | 90 (21) | 128 (19) |

\*Differential number of missing values across variables
Abbreviations: High income countries (HIC); Low and middle income countries (LMIC)

Among respondents from LMICs (n=263), half reported receiving updated guidelines for the provision of maternal and newborn care reflecting measures for the COVID-19 outbreak, compared to 82% of those from HICs (Table 2). Some LMIC-based respondents (particularly from Tanzania, Rwanda, Uganda, and India) expressed their concern over the lack of updated guidelines and protocols. An obstetrician/gynaecologist from Uganda remarked: “*I am worried that no national guidelines [are] rolled out yet in regards to care for pregnant women and newborns*.” Some midwives working in HICs requested clearer guidelines on the provision of midwifery care during home visits. Nearly all respondents reported having searched personally for information on COVID-19 (92%), and received informal guidance from colleagues (90%, Table 2). Nonetheless, some participants in LMICs were worried about the lack of access to/availability of evidence on the effects of COVID-19 during pregnancy and the possibility of in-utero transmission, and transmission through breast milk to newborns. More than half (61%) of respondents perceived that patients’ questions regarding COVID-19 were being adequately answered by healthcare providers in their respective facilities. However, only 19% of participants felt that they were completely knowledgeable of the measures that should be taken to provide care to COVID-19 maternity patients (Table 2). Almost half of respondents indicated that their facilities had shared materials with maternity patients on COVID-19, and these materials were mainly disseminated through health facility websites, leaflets/fliers, posters, and on social media.

Some participants from LMICs such as India, Bangladesh, Bolivia, and Syria expressed concerns regarding patients’ degree of application of instructions, particularly those related to social/physical distancing and hygiene measures. An obstetrician/gynaecologist from India mentioned worrying about *“patients and relatives not following instructions given by staff members”*. As described by a nurse from Syria, some respondents attributed this to a *“lack of awareness and knowledge, and indifference among beneficiaries”*. Furthermore, the implication of the local communities and sharing responsibilities in terms of application of the hygiene and social/physical distancing measures were mentioned by some respondents. A midwife from Bolivia worried *“that not enough is being done on a personal level by patients to keep themselves safe”*.

### Work environment adaptations in response to COVID-19

Three quarters of participants from HICs reported that their facilities had set up a well sign-posted general entrance and screening area for COVID-19 suspected cases, compared to 37% of respondents from LMICs. Among HIC respondents, 83% reported that their facilities reserved isolation rooms for suspected COVID-19 cases, compared to 57% of LMIC respondents (Table 3). The majority of respondents (62%) reported that their facilities have designated a COVID-19 liaison person or team. These teams were most commonly assigned at the level of the facility (56%), followed by both at the levels of facility and maternity ward (27%), and in the maternity ward alone (17%). Screening for COVID-19 symptoms among maternity patients was also more commonly reported by respondents working in HICs (76%) versus 47% in LMICs. For example, antenatal care (ANC) patients - both outpatients and inpatients - were screened either in person or over the phone before scheduling appointments. The ability to order COVID-19 tests for maternity patients was available for 61% of respondents in HICs; but limited in LMICs (23%), rural areas (9% in LMICs and 28% in HICs) and completely unavailable to respondents working in refugee and/or displaced persons camps (n=6, data not shown).

**Table 3.**
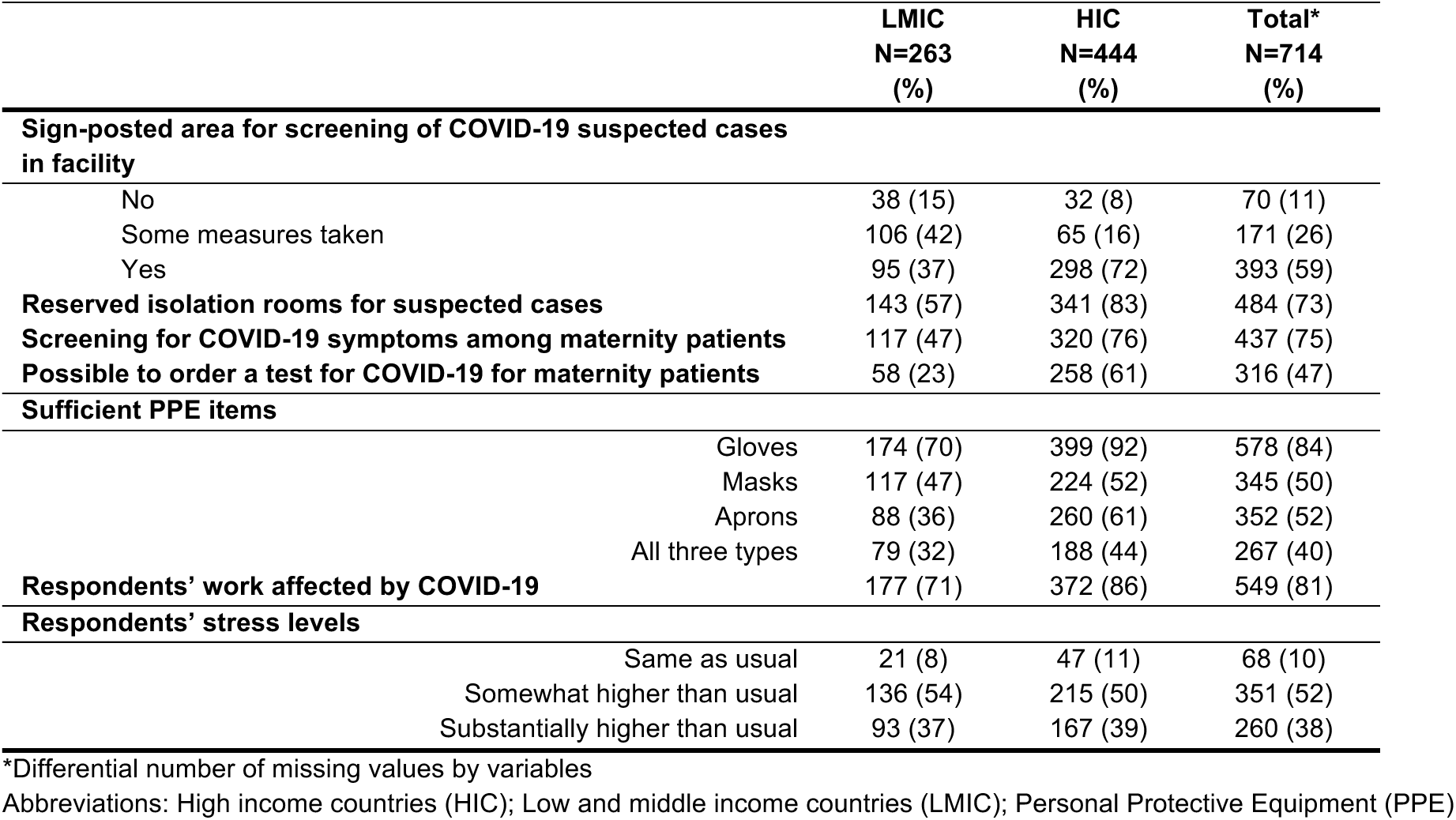
- Response to COVID-19 among maternal and newborn health professionals and their workplaces, by country income category

The lack of COVID-19 symptom screening and the inability to order tests constituted major concerns for respondents, who perceived these deficiencies as threats to the safety of the workforce and of other patients. A midwife from Canada wrote, *“I’m worried about being infected by someone who is asymptomatic, and then being a vector to others.”* Compromising patients’ and healthcare providers’ safety was also viewed as stemming from deficiencies in availability of PPE, including face masks, gloves, and aprons. These findings were consistent across all settings but more prominent in LMICs (Table 3). A midwife from the United Kingdom pleaded, *“Let midwives who are in close contact with women wear masks. […] Please let us use masks for all.”* Additionally, respondents advocated for clear guidelines and unified protocols regarding the appropriate use of PPE. For example, a nurse-midwife from the United States wrote, “[…] *as of now we are not allowed to wear masks and goggles unless delivering a patient, we’re told to « <u>take the mask off or go home</u>* » *that we’re scaring the patients.”* Despite the need to feel protected, wearing additional PPE can be burdensome. It was described as time-consuming and respondents worried about resulting delays in the provision of emergency care because of having to don and doff PPE. Additionally, they were concerned that PPE might reduce their ability to communicate clearly with patients, such as a midwife from Denmark who remarked that *“[i]t can be hard to connect with people through masks and [goggles] (facial expressions are harder to read).”*

The majority (81%) of respondents noted that their work had been affected by the COVID-19 outbreak and that their stress levels were either somewhat or substantially higher than usual (90%, Table 3). In the words of an obstetrician from Mozambique: *“My stress level at this point is immeasurable. Every time a pregnant woman with flu-like symptoms [visits the health facility], I feel almost completely lost and I end up only of [thinking about this] patient. I need to be equally protected and I don’t feel any protection from whoever [is responsible for protecting me].”* A major challenge reported was the decrease in skill mix and shortages of qualified staff, either because of symptoms, self-isolation after potential exposure, or not being able to get to their workplace due to lockdowns and transport restrictions as described by a midwife in Uganda: *“[t]ransport to work is a big challenge due to lockdown; many staff live far away from the hospital. The staff who manage to come to work hurry to leave the hospital early to observe the curfew time of 7.00* p.m.” This shortage has led to an increase in the workload and unexpected changes in work schedules. Anxiety and exhaustion levels have increased because of these rapid changes, and some respondents expressed the need for more support from management. In certain contexts, healthcare facilities were increasingly relying on locum workers and students to fill staffing shortages. An obstetrician/gynaecologist who headed a department in Uganda reported that *“[t]here is no more clear work schedules as I get to attend many unscheduled/emergency meetings […]. The staff are very anxious and panicky and need talking to all the time, which is exhausting.”*

### Changes to the care provided to women and newborns

We analysed responses from 397 health professionals who completed the optional module. Figure 1 shows the main reported changes in service provision and utilisation, care content and quality, and care process adaptations across the continuum of maternal and newborn care. In all settings and across the continuum of care, participants reported seeing fewer patients at healthcare facilities. This was described as the result of transportation difficulties in accessing health facilities or due to women’s fear of contracting COVID-19 at the facility. A nurse-midwife from Kenya wrote, *“[a]ccessing inpatient antenatal care [is] minimal. Women fear to [get] infected with COVID-19 if [they are present] in hospitals. Most of them keep off from hospital even when they are sick’*. Most respondents noted that their facilities have shortened visiting hours and reduced the number of visitors allowed, while others are screening visitors for symptoms, or have banned visits altogether. Importantly for the support of women during labour and childbirth, facilities are reportedly limiting the number of labour companions to one person designated as the single visitor allowed to stay with the mother after birth or banning birth companions altogether. This raised concerns among healthcare providers regarding the reduced support available to women, and increased workload on the staff. An obstetrician from the Czech Republic remarked that: *“[the] Gynaecological and Obstetrical Society has recommended to ban partners and doulas from accompanying a woman at birth - outrageous!!!”*

**Figure 1.**
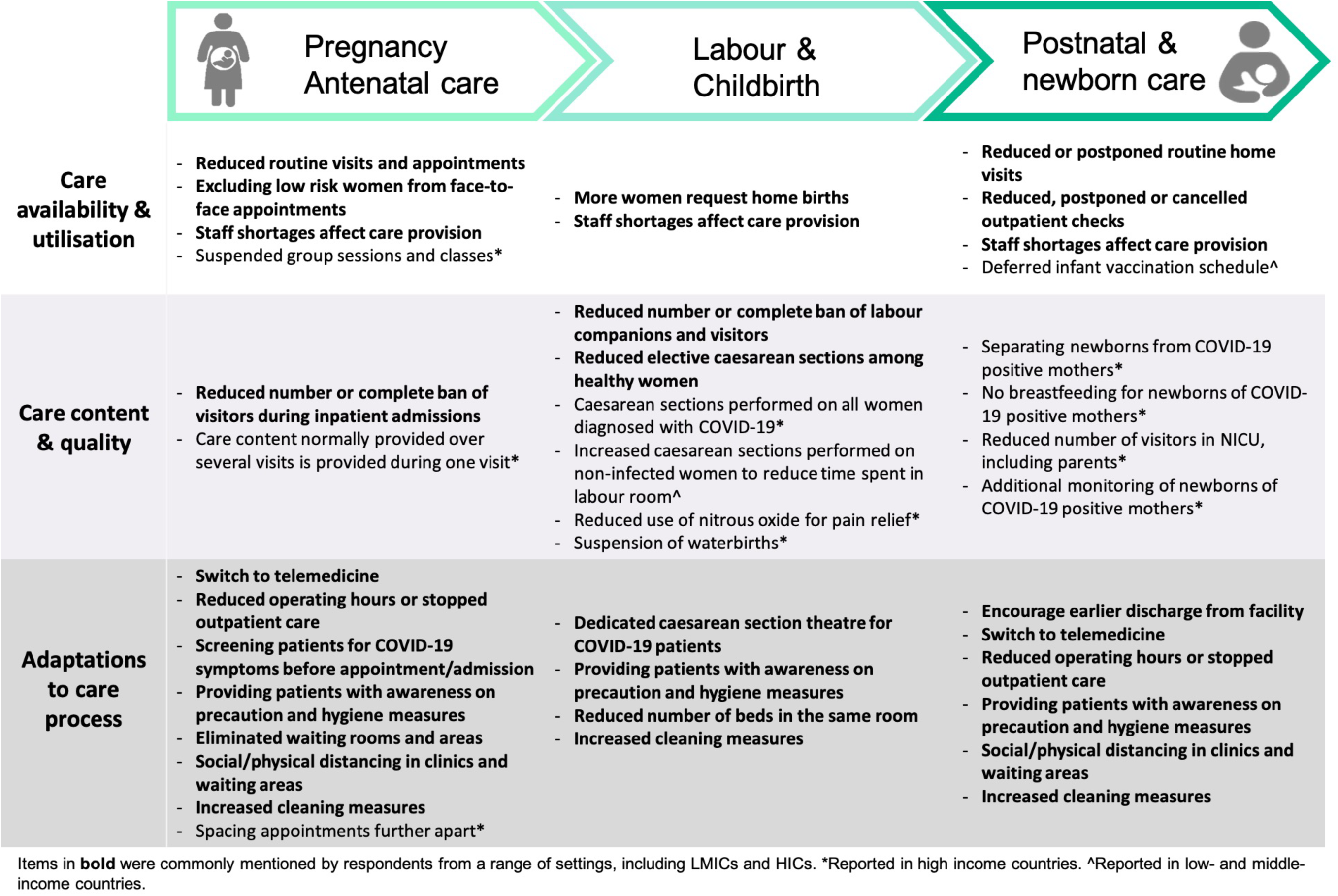
Reported changes to service provision across the continuum of maternal and newborn care

Some healthcare facilities were implementing social/physical distancing measures in the waiting areas of the outpatient departments, and in hospital rooms by reducing the number of beds. Yet, this recommendation is challenging to meet in facilities where resources are limited - an obstetrician/gynaecologist from India noted that “[it is] *not practically possible [to place each patient in a separate birthing room] in our set up”*. Non-essential services including elective gynaecological procedures and infertility treatments were being postponed or cancelled. In several settings, responding to the COVID-19 outbreak affected the delivery of routine ANC, which became restricted to the management of high-risk patients. A respondent from New York reported a *“significant decrease in number of ANC visits”*, whereby new policies recommend the reduction of the number of face-to-face visits during pregnancy *“from 10-12 to four”*. Other changes include eliminating the waiting area, spacing face-to-face appointments to reduce contact between patients, and cancelling all group activities such as health education sessions or group counselling.

Respondents also reported a shift to telemedicine for the provision of both antenatal and postnatal care (PNC), including breastfeeding counselling. Although telemedicine was considered a priority in certain LMICs where it is not implemented yet, participants acknowledged the challenges associated with this service provision modality. This includes lack of access to adequate communication infrastructure among women. Respondents from both LMICs and HICs noted that the demand for home births has increased and that new practices aimed to reduce induction of labour. In certain HICs, induction of labour was reported to be discouraged before 41 weeks of gestation. Changes in pain relief options for labouring women in HICs included decrease in the use of nitrous oxide to reduce the risk of infection transmission through aerosols, and suspended waterbirths. Across all settings, caesarean sections were reported as a commonly performed procedure among women who were diagnosed with COVID-19. Some respondents noted that their facilities have dedicated operating theatres specifically for this purpose. On the other hand, the numbers of elective caesarean sections have reportedly decreased among “healthy” maternity patients. However, this was not consistent in facilities where certain efforts were made to reduce the duration of labour and the time spent in the labour room by augmentation. As a result, respondents speculated about a potential rise in caesarean section rates in their facilities, as noted by an obstetrician/gynaecologist from India: “*We will not allow as much time in second stage [of labour], […] this is likely to push up our caesarean rate.”*

Respondents frequently mentioned shortened length of stay in facilities after childbirth; for example a reduction *“to 6-8 hours from 24 [or more hours]”* (midwife from Canada). This was worrisome for some respondents as noted by a midwife from the UK: *“[the] lack of time and staff will lead to mothers and babies going home with very little feeding support or knowledge which will have a short and long term impact on their health and ability to deal with infections.”* Routine postnatal checks are being postponed in certain cases or substituted with telemedicine. A nurse-midwife from the United States reported that *“[w]e are postponing the routine postpartum visit until 12 weeks postpartum, and are prescribing most contraceptives over the phone or […] and breastfeeding support is all done virtually.”* Changes to newborn postnatal care were infrequently reported, and mainly included monitoring and isolation of babies of mothers with confirmed COVID-19. Three respondents from India noted that the infant vaccination schedule was disrupted or postponed. Overall, respondents expressed their concern over the uncertain impact of reduced face-to-face interactions on the quality of care. A midwife from the UK wrote: *“[w]hilst I completely see the need to restrict our face-to-face care to protect staff and patients, my heart just breaks for women and families who we won’t be able to offer the full range of midwifery support to… i.e. BF support, daily visits, and just generally our time”*. Maternal and newborn health professionals feared that the changes to the standards of care would lead to poor health outcomes among women and newborns and, subsequently to the loss of progress achieved in certain indicators (e.g., stillbirth rates). “*I am also worried about the implications of the policies that call for separating newborns from COVID-19 positive mothers immediately after birth, without allowing for skin-to-skin or delayed cord clamping,”* wrote a nurse-midwife from the United States.

## Discussion

This paper uses a rapid collection of data from health professionals providing care to women and their babies globally. We describe preparedness for COVID-19, response to COVID-19, personal experience in the workplace, and changes in provision of care and care processes.

### Preparedness

We found that respondents actively sought information related to COVID-19 through personal searches and existing informal networks. Studies show that healthcare providers commonly resort to such sources to fulfill information needs.^33^ Knowledge gaps were generally related to the impact of COVID-19 on pregnancy and health outcomes for the mother and newborn, or to guidance on the management of COVID-19 maternity cases. There is a high possibility that unreliable information related to the outbreak might be accessed, particularly on social media platforms.^34^ Facility-specific creation and distribution of guidelines for managing maternity patients is somewhat lagging behind despite frequent general updates published by Ministries of Health and professional associations.^35–41^ Information sharing channels must be established to secure providers’ timely access to accurate information that empowers them to respond to patients’ needs.^42–44^ Midwives supporting pregnant and labouring women during the pandemic,^45,46^ and particularly those who are practicing independently, have voiced the need to access clear guidelines for providing care during home visits.^47^

### Response

Our results highlight variability in the facility-level response to COVID-19 between HICs and LMICs, including sharp differences in updating guidelines, setting-up signage and patient/visitor screening, testing availability, and dedicating isolation rooms for maternity patients with confirmed or suspected COVID-19. These discrepancies could stem from the differential progression of the outbreak (whereby more respondents from HICs reported having provided care to COVID-19 confirmed or suspected maternity cases than those from LMICs). These differences could also be partly attributed to the limited capacities and resources of healthcare systems in some LMICs.^48^ There is speculation that the outbreak in African countries might be attenuated, but equally possible that trends similar to those witnessed in Europe might be observed.^49,50^ This indicates an urgent need to mobilise resources in resource-limited settings, improve testing capacities, and upgrade the responses, including at maternity facilities. The total absence of testing for suspected patients in refugee and/or displaced persons camps reported by all respondents with such experience raises concerns. Living conditions in these under-served settlements, such as overcrowding and lack of adequate water and sanitation, make the implementation of basic infection prevention and control measures nearly impossible.^50–53^ Displaced women and their newborns face sub-optimal access to ANC, skilled attendance at birth, PNC, and vaccination, and subsequently experienced poor health outcomes even prior to the pandemic-induced disruptions of essential services.^54–57^ Global and local efforts must be established to ensure that displaced populations have access to appropriate infection prevention measures, testing and treatment, and to quality maternal and newborn services to halt anticipated exacerbations of negative health outcomes.^29,52^

### Personal Experiences

Consistently with experiences from previous infectious disease outbreaks and emergencies, healthcare workers providing essential services to women and newborns during this pandemic experience increased levels of stress and anxiety.^20,44^ Stress levels in LMICs were comparable to those in HICs even though countries were battling different stages of the outbreak. This might be due to uniformly reported changes in working hours, inability to reach health facilities, and shortages in skilled workforce (some of which were attributable to lockdowns and other blanket measures to combat the spread of COVID-19) leading to higher workloads, which can lead to staff burnout.^48,58^ Wilson et al. compiled a list of measures that could prevent burnout among maternity care providers.^44^ Other lessons learned from past epidemics include the provision of emotional, social and mental health support to care providers, and ensuring that adequate levels of support is available to them from facility management.^20,59^ As our findings show, this can create additional burdens to management and special efforts should be placed to provide support to this group.^44^ With the increasing reliance on students and trainees to compensate for staff shortage, Wilson et al. consider this group to be more exposed to stressors considering their lack of experience, and therefore senior colleagues should actively advocate for their wellbeing.^44^ Future research should explore the availability of mental and social support to maternal and newborn healthcare providers during the pandemic, and its effectiveness.^60^

Another cause for increased stress levels among providers is the fear for their own/their relatives’ safety, in addition to the safety of their patients, which is intensified by inadequate access to PPE. One reason for that is that in some facilities, PPE supplies are prioritised for departments treating COVID-19 cases and not reaching maternity wards, which is common in vulnerable settings. Workers who provide essential maternity care and their patients, could thus experience uneven risks of nosocomial infection during outbreaks.^61,62^ In some countries, obstetricians/gynaecologists commonly work in multiple facilities across the public and private sectors, and their risk of exposure might be exacerbated by the higher number of contacts (other healthcare workers and patients) they experience in this dual practice.^63^ Although PPE items are essential, the WHO issued guidelines that promote their rational use given universal shortage.^64^ The application of these guidelines must be unified within healthcare facilities, and clearly communicated to maternity and newborn healthcare providers and explained to patients.^44^ Health workers caring for women around the time of birth might be used to wearing PPE, however it can make them feel dehumanized, and the donning and doffing PPE is time-consuming and might delay the provision of emergency services.^20,21^

Healthcare providers also worry about the consequences of rapidly changing practices and the uncertainty of their impact on health outcomes. The perceived changes in healthcare seeking behaviours include fewer visits to the healthcare facilities, shortened lengths-of-stay after childbirth, less access to adequate ANC, and in certain cases, disrupted immunisation schedules. Our findings support the narratives told by healthcare providers regarding the discontinuity of basic services,^65^ and align with disruptions witnessed during previous infectious disease outbreaks.^58,66–68^ These disruptions have previously led to an increase in maternal and neonatal mortality,^22,69^ and currently there are signs of similar trends reported from two maternity hospitals in Uganda.^70^ Our knowledge of the impact of these changes is restricted to predictions resulting from modelling which strongly suggest a threat to achieved improvements in LMICs.^28,29^ The actual impact is yet to be quantified^28^ and the effect of these changes in HICs remains unclear. Prioritised measures depending on contextual needs must be put in place to mitigate these indirect consequences of the pandemic.^28,30^

Although some of the changes to care content and process are consistent with the updated guidelines on essential care provision,^40,71–73^ other modifications diverge from available evidence, and could ultimately reverse achieved progress if proper action is not taken. These include the elimination of birth companions altogether,^73,74^ reducing or banning visitors to maternity wards, performing caesarean section on all COVID-19 positive women,^74,75^ augmenting labour or performing unindicated caesarean sections to gain control over timing of deliveries, separating newborns from COVID-19 positive mothers including not allowing breastfeeding,^76^ and drastically reducing length-of-stay after facility birth with fewer home visit follow-ups.^47^

These practices could deny women’s access to quality, respectful intrapartum and postpartum care, and jeopardize their wellbeing and that of their babies.^77^ Unlike curative health services, maternity care focuses on providing holistic support to women going through a normal physiological process; both over and under-intervention can result in massive preventable burden. Another alarming adaptation to COVID-19 is freezing or postponing routine immunisation schedules.^78^ Temporary disruptions to routine immunisation were only reported in India in our survey, however other LMICs have implemented similar measures.^78^ This can result in overall declines in population coverage, and catch-up campaigns should be prioritised following the relaxation of preventive measures to ensure the sustainability of achievements.^78,79^ The introduction of new models of care such as telehealth guidance^29,71^ was described as a ‘virtually perfect solution’ to ensure sustained care provision during the pandemic.^80^ Yet this model’s feasibility is not universal to all healthcare services. Midwives dread this mode’s disturbance of the quality of provided care,^47^ and providers in LMICs consider this as an added barrier to achieving equitable access to essential services for women and families who lack the needed resources.

As several qualitative and ethnographic studies have shown,^81,82^ healthcare seeking behaviours of patients and local communities relies, among others, on the provider-patient relationship and common cultural, economic and social understanding of health and hygiene.^83,84^ Moreover, hierarchical issues such as racial and social discrimination may have a significant impact on the quality of maternal and newborn healthcare, as it has already been highlighted in, for example, West-African urban areas and Malagasy hospitals ^81,82,85^, to only cite a few. Dynamics of mutual incomprehension between patients and providers about the attitudes toward and measures necessitated by the outbreak may be taken into account regarding the impact of COVID-19 on maternal and newborn healthcare; especially (but not only) concerning regions and countries where there is a strong medical and cultural pluralism towards the healthcare seeking behaviours. Although only a few respondents mentioned some resistance of patients and local communities to increased hygiene and physical distancing measures in facilities, we know from previous outbreaks such as Ebola that understanding social and cultural responses to the epidemics is essential to prevent healthcare disasters.^86^ Furthermore, many respondents addressed the reduction of the number of visitors allowed during labor and childbirth, when such measures do not seem justified. Limiting social support during maternal and neonatal care can put mother and newborn at risk, more particularly in healthcare facilities with an ordinarily high lack of staff and irregular drug delivery where families and surrounding play a crucial role in limiting the impact of these shortcomings.^82,87^ A local understanding of healthcare seeking behaviors and social maternal health organization must take place to avoid a top-down management of the outbreak guidelines that may miss the mark of these local pre-existing factors.^88^

## Limitations

We acknowledge that lack of representativeness and related sample bias are limitations of this sampling approach. Our sample might over-represent higher qualified cadres of health professionals in settings with limited use of technology among lower cadres of staff, and under-represent staff who are most overstretched in providing healthcare, or those with limited or no access to internet connection, as we received few responses from professionals working in lower-level facilities, particularly in LMICs. The representativeness of the sample is affected by the availability of the survey in three languages (English, French, and Arabic) for a longer time than the remaining nine languages. Additionally, some cadres, such as neonatologists and paediatricians, were less represented. The questionnaire asks about facilities where respondents work, which is not relevant to independently practicing professionals, especially midwives; this might have discouraged some of them from completing the survey. Finally, data were collected across countries that were going through different stages of the outbreak. As previously mentioned, this could account for country-level discrepancies in the response, and some of the differences seen between HIC and LMIC respondents. We intend to address some of these limitations in survey rounds.

## Conclusion

Despite the limitations, this is the first study attempting to describe the preparedness for, response to and effect of, the COVID-19 pandemic on the provision of maternal and newborn care. The multi-country aspect of the survey allows for a one-stop platform for lessons to be learnt and shared across systems. Our findings, ideally combined with an understanding of women’s perspectives, hold enormous potential for creating a timely and evidence-based decision-making platform. Disseminating health workers’ voices to planners, programmers and policymakers is crucial to guide the development of global and contextual guidelines for practice and preparedness.

The COVID-19 pandemic illustrates the worldwide susceptibility to emergencies, which is not restricted to healthcare systems in LMICs. Responding to this crisis is proving to be challenging for health systems and providers, and affects access to basic health services across the globe. Preparedness for the global pandemic might have been equally inadequate for health systems in LMICs and HICs in some aspects, such as shortage in skilled staff, providing training and simulations, and PPE sufficiency. However, it is likely that HICs were able to respond more effectively, due to better health system resilience such as existing coordination systems to develop and implement changes to protocols.^89^ Findings from this study will be useful in supporting the development of effective responses to the main issues identified, both during the COVID-19 pandemic and more broadly during future health system shocks.

## Data Availability

Anonymised data analysed during the current study will be made available from the corresponding author upon reasonable request.

## Acknowledgments

We would like to thank the study participants who took time to respond to this survey despite the difficult circumstances and increased workload. We acknowledge the Institutional Review Committee at the Institute of Tropical Medicine for providing helpful suggestions on this study protocol, and for the expedited review of this study. We thank all study collaborators and colleagues who distributed the invitation for this survey and provided suggestions on the questionnaire, including the co-authors of this paper, Dr Susannah Woodd and Dr Jean-Paul Dossou. We are immensely grateful to all those who volunteered to translate the survey, including Dr Francesca L Cavallaro, Dr Bouchra Assarag, Dr Karima Khalil, Dr Jose Penalvo, Dr Sonia Menon, Dr Claudia Nieto Sanchez, Virginia Castellano Pleguezelo, Dr Werner Soors, Verónica Blanco Gutiérrez, Dr Raffaella Ravinetto, Dr Manuela Straneo, Francesca Palestra, Dr Stacy Saha, Dr Shirajum Munira, Dr Mitsuaki Matsui, Wnurinham Silva, Dr Leonardo Chavane, Dr Enny Cruz, Min Dai, Dr Li Na, Dr Marc Snel, Dr Anna Larionova, Dr Vladimir Gordeev, Dr Christiane Schwarz, Evelyn Lesta, Elibariki Mkumbo, Dr Amani Kikula, Dr Elly Mertens, and Hilde Cortier.

## Author contribution

LB conceptualised the study and obtained funding. All authors contributed to the design of the study and development of the survey tool. AS analysed the data. CA, LB, EH and AS wrote the original draft of the manuscript. All authors contributed to the development of the manuscript, and read and approved the final version. The corresponding author attests that all listed authors meet authorship criteria and that no others meeting the criteria have been omitted. AS is the guarantor.

## Ethical approval

This study was approved by the Institutional Review Committee at the Institute of Tropical Medicine (Antwerp, Belgium) on March 20, 2020 (approval reference 1372/20).

## Patient and Public Involvement

No patient or public involvement took place in the design or conduct of this study. We involved maternal and newborn health professionals, experts in health systems, infectious diseases, infection prevention and control, and maternal health epidemiologists, and public health researchers from various global settings in the design of this study and the survey tool.

## Dissemination to participants and related communities

The authors intend to disseminate this research through social media, press releases, and media departments and websites of authors’ institutions.

## Transparency

The guarantor affirms that this manuscript is an honest, accurate, and transparent account of the study being reported; that no important aspects of the study have been omitted; and that any discrepancies from the study as planned (and, if relevant, registered) have been explained.

---

**Supplementary File 1.**
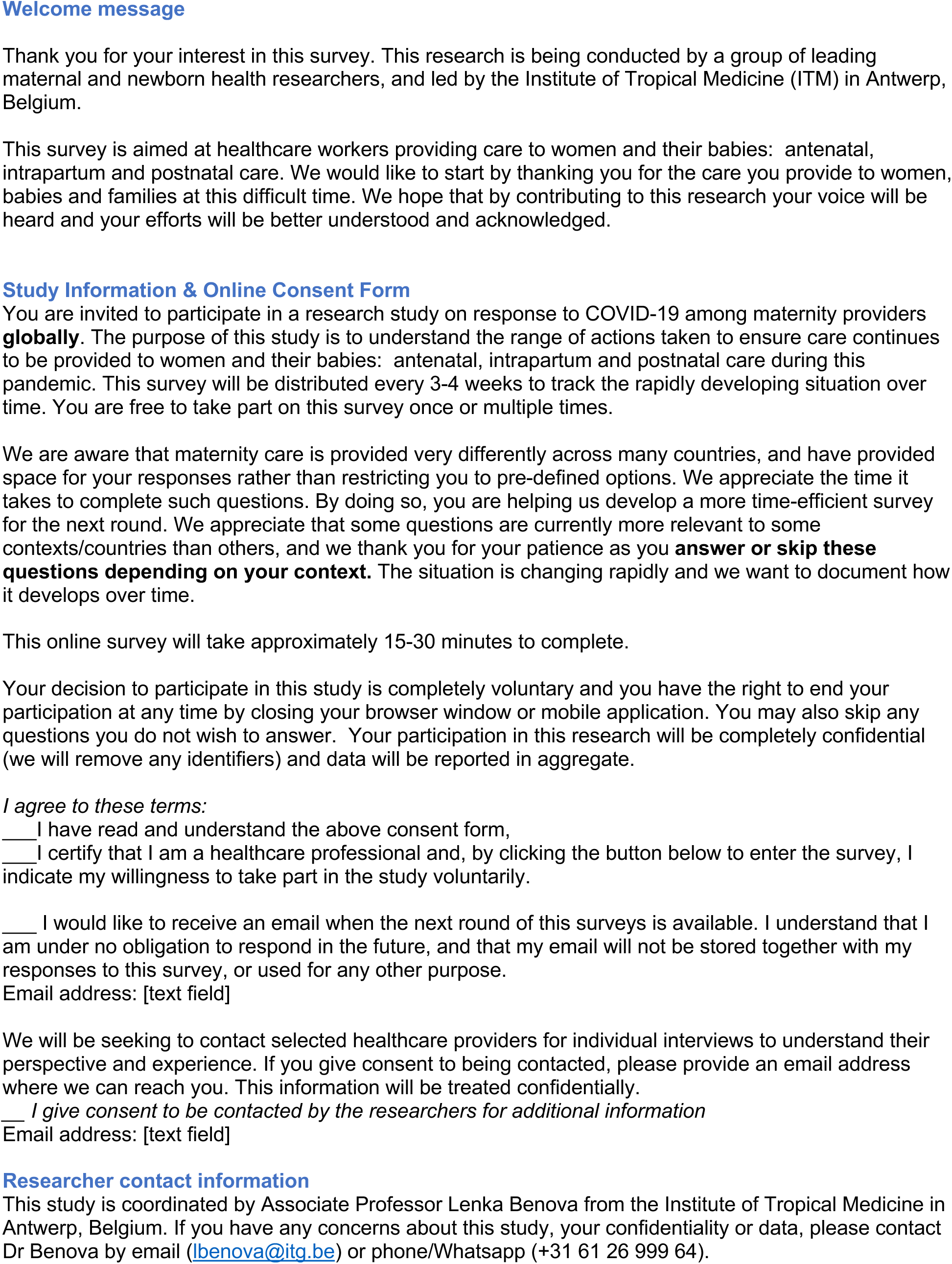
- Online Questionnaire – Round 1.

**Part 1.** We would like to ask a few questions about your background.

**Part 1. We would like to ask a few questions about your background**
| Q# | Question | Response |
| --- | --- | --- |
| 1 | In which **country** are you based (providing healthcare) at the moment? | [drop down menu of countries] |
| 2 | In which **region** of the country do you work?<br>(please provide region, district, province, state, or governorate) | [free text] |
| 3 | What is your **job**?<br>(choose one) | - Midwife<br>- Nurse-midwife<br>- Nurse<br>- Obstetrician/gynecologist<br>- Anaesthesiologist<br>- Surgeon<br>- Neonatologist<br>- Pediatrician<br>- General practitioner<br>- Medical doctor (other/no specialisation)<br>- Medical student/intern/resident<br>- Medical officer<br>- Clinical officer<br>- Nurse or midwife in training<br>- Ultrasound technician/ sonographer<br>- Health technician<br>- Paramedic<br>- Community health worker/ Outreach worker<br>- Lactation counsellor<br>- Other: specify |
| 4 | What is your **position**?<br>(choose one) | - Head of facility (director, administrator)<br>- Head of department or ward<br>- Head of team<br>- Team member<br>- Locum or interim member<br>- Other: specify |
| 5 | What is your **gender**?<br>(choose one) | - Female<br>- Male<br>- Other/Prefer not to say |
| 6 | What **type of maternal and/or neonatal health care** do you currently provide as an individual?<br>(mark all that apply) | - Outpatient antenatal care<br>- Outpatient (home-based) childbirth care<br>- Outpatient postnatal care<br>- Outpatient breastfeeding support<br>- Inpatient antenatal care<br>- Inpatient childbirth care<br>- Inpatient postnatal care (mother and/or babies)<br>- Surgical care<br>- Neonatal care for small and sick newborns<br>- Home visits<br>- Community outreach, home visits, health education outside facility<br>- Abortion care<br>- Post-abortion care<br>- Other: specify |

**Part 2.** Setting: Can you tell us about the facility setting in which you work now.

| Q# | Question | Response | Notes |
| --- | --- | --- | --- |
| 1 | In which **level** of health care institution do you primarily work?<br>(if none of the response options fit well, please use the "Other" option and write what your facility type is called in your country) | - Referral hospital<br>- District/regional hospital<br>- Health center<br>- Polyclinic<br>- Clinic<br>- Health post/unit<br>- Dispensary<br>- Other: specify |  |
| 2 | What organisation **type** is your institution? | - Public (national)<br>- Public (university or teaching)<br>- Public (district level or below)<br>- Social security<br>- Health insurance or HMO<br>- Private university<br>- Private for profit<br>- Non-governmental<br>- Faith-based or mission<br>- Other: specify |  |
| 3 | In what type of **geographic area** is your facility located? | - Large city (>1 mil inhabitants)<br>- Small city (100,000 to 1 mil inhabitants)<br>- Town (<100,000 inhabitants)<br>- Village or rural area<br>- Refugee or displaced persons camp<br>- Other |  |
| 4 | How many **maternity beds** does your facility have (include antenatal, labour/childbirth and postnatal). Approximate number is ok | None<br>Number: _____[free text] |  |
| 5 | How many **births** took place in your facility in **2019**?<br>Approximate number is ok | None<br>Number: _____[free text]<br>Don't know |  |
| 6 | Does your facility provide **caesarean sections**? | Yes<br>No | If no, skip to 8 |
| 7 | If yes, what is the **csection rate** (% of births by csection) in your facility? Approximate number is ok | Number: _____[free text]<br>Don't know |  |
| 8 | Does your facility have an **Intensive Care Unit (ICU)** which can admit **women** with obstetric complications? (ICU is defined as a clinical area where ventilatory support can be provided) | Yes<br>No<br>Don't know |  |
| 9 | Does your facility have a **neonatal intensive care unit (NICU)**?<br>(Neonatal intensive care is defined as a unit that provides invasive ventilatory support to small and sick newborns, not just CPAP) | Yes<br>No<br>Don't know |  |
| 10 | Does your facility **receive maternity referrals** from other facilities, meaning that patients are sent to your facility from other health facilities? | Yes<br>No<br>Don't know |  |
| 11 | Is running **water and soap** always available for hand hygiene on your ward **for the use of staff**? | Yes<br>No<br>Don't know |  |
| 12 | Is running **water and soap** always available on your ward for the use of **patients, visitors, companions**? | Yes<br>No<br>Don't know |  |
| 13 | Is there always **sufficient water and disinfectant for cleaning surfaces**? | Yes<br>No<br>Don't know |  |

**Part 3.** COVID-19 preparedness.

Part 3. COVID-19 preparedness
| Q# | Question | Response | Notes |
| --- | --- | --- | --- |
| 1 | Has your institution or ward provided you with **any information** on how to prepare for COVID-19? | Yes<br>No | No – skip to 6 |
| 2 | What did you learn from this information? Please list main areas or themes | [free text] |  |
|  | On a scale from 1 (poor) to 5 (excellent), how would you rate the following dimensions of this information: | 1 – poor<br>2 – somewhat useful<br>3 – average<br>4 – good<br>5 – excellent<br>Not applicable |  |
| 3 | Clarity |  |  |
| 4 | Helpfulness for your daily work |  |  |
| 5 | Value in helping you feel safe |  |  |
| 6 | Has your institution or ward provided you with any training on COVID-19, for example simulations or drills? | Yes<br>No |  |
| 7 | Have you received new or updated guidelines specifically for the provision of care to pregnant, labouring or postpartum women and their newborns because of COVID-19? | Yes<br>No | If no, skip to 9 |
| 8 | If yes, which guidelines? Please list all used (WHO, FIGO, ICM, RCOG, RCPCH, RCN, RCM, COINN, your country's Ministry of Health, Country professional organisations etc) | Free text |  |
| 9 | Have you personally searched for source of guidance and sources of information to prepare for COVID-19 in your work? | Yes<br>No |  |
| 10 | Have you received information related to COVID-19 and your work informally through other colleagues (in your own facility or outside)? | Yes<br>No |  |
| 11 | Have you been a part of any self-organisation on the part of healthcare workers in response to the COVID-19 outbreak? (exchange of information, virtual discussion groups (Whatsapp, Facebook, etc) | Yes<br>No |  |
| 12 | Has your facility published or distributed any materials (brochure, flier, posters, etc) covering COVID-19 targeted toward pregnant, labouring, or postnatal women? | Yes<br>No<br>Don't know | If no or don't know - skip to 15 |
| 13 | What kind of information is on these materials? (social distancing, symptoms, when/how to self-isolate, when/where to seek care etc.) | Free text |  |
| 14 | In what form is it provided? (mark all that apply) | -Health talks<br>-Leaflets/fliers<br>-Posters<br>-Counselling during consultations<br>-Facility website<br>-Phone line with advice<br>-Other: specify |  |
| 15 | In your facility, do you feel that patients' questions about COVID-19 are being addressed adequately by staff? | Yes<br>No<br>Don't know |  |
| 16 | What are you worried about most at this time in regard to **being able to provide care to women and newborns**? | Free text |  |
| 17 | Has your facility set up a well sign-posted general entrance and screening area for COVID-19 suspected cases? (regardless whether for maternity patients or not) | -Yes<br>-Some measures taken but not done well<br>-No measures taken<br>-Don't know |  |
| 18 | Has your facility reserved **isolation rooms** for COVID-19 suspected cases? | Yes<br>No<br>Don't know |  |
| 19 | Has **routine cleaning of the maternity ward** changed in response to COVID-19? | Yes, increased<br>Yes, decreased<br>Unchanged<br>Don't know |  |

**Part 4.** Response to COVID-19 in your facility.

Part 4. Response to COVID-19 in your facility
| Q# | Question | Response | Notes |
| --- | --- | --- | --- |
| 1 | Is your facility currently screening for COVID-19 symptoms among maternity patients? | Yes<br>No<br>Don't know<br>Other: specify |  |
| 2 | Is it possible to order a test for COVID-19 at the moment for maternity patients at your facility? | Yes<br>No<br>Don't know<br>Other: specify | If no/DK: skip to 4 |
| 3 | If yes, how long does it take to get a result? (note whether your response is in hours or days) | [free text] |  |
| 4 | Are the testing criteria for COVID-19 clear to you? (the conditions/symptoms for which a test can be ordered) | Yes<br>No | If no – go to 6 |
| 5 | Can you describe these criteria | [free text] |  |
| 6 | Have you had any maternity patients with COVID-19 in your facility so far? | Yes, suspected<br>Yes, confirmed<br>Yes, both confirmed and suspected cases<br>No<br>Don't know<br>Other: specify | If no or DK – go to 8 |
| 7 | If yes, approximate number | [free text] |  |
| 8 | Is there a designated COVID-19 lead person / liaison or team in the maternity ward or the facility? | -Yes, in maternity<br>-Yes, in facility as a whole<br>-Yes, both in the maternity ward and in the facility as a whole<br>-No, neither maternity nor facility<br>-Don't know |  |
| 9 | Were you aware that the WHO developed a pregnancy/postpartum module to be included in the current Case Report Forms (CRFs) recommended to report COVID-19? cases (https://isaric.tghn.org/novel-coronavirus/) | Yes<br>No | If no, skip to 11 |
| 10 | If yes, are you using this module or preparing to use this module in your facility? | Yes, already using<br>Yes, preparing to start<br>No<br>Don't know<br>Other: specify |  |
| 11 | On a scale from 1 (not at all) to 5 (I am very clear), do you personally feel you know what you should do if a woman with COVID-19 symptoms arrives in your facility today? | 1 – Not at all clear<br>2 – Some points are clear to me, but I am not confident in what to do<br>3 – Somewhat clear but major issues remain<br>4 – I am mostly clear but some questions / areas of concern remain<br>5 – I am very clear |  |
| 12 | On a scale from 1 (not at all) to 5 (completely), do you feel that you are | 1 – not at all<br>2 – minimal protection |  |
|  | sufficiently protected from infection with COVID-19 in your workplace? | 3 – some protection<br>4 – well protected<br>5 – completely protected |  |
| **Is a sufficient quantity of personal protective equipment (PPE) available to you?** |  |  |  |
| 13 | Gloves | Yes - No |  |
| 14 | Masks | Yes - No |  |
| 15 | Aprons | Yes - No |  |

**Part 5.**
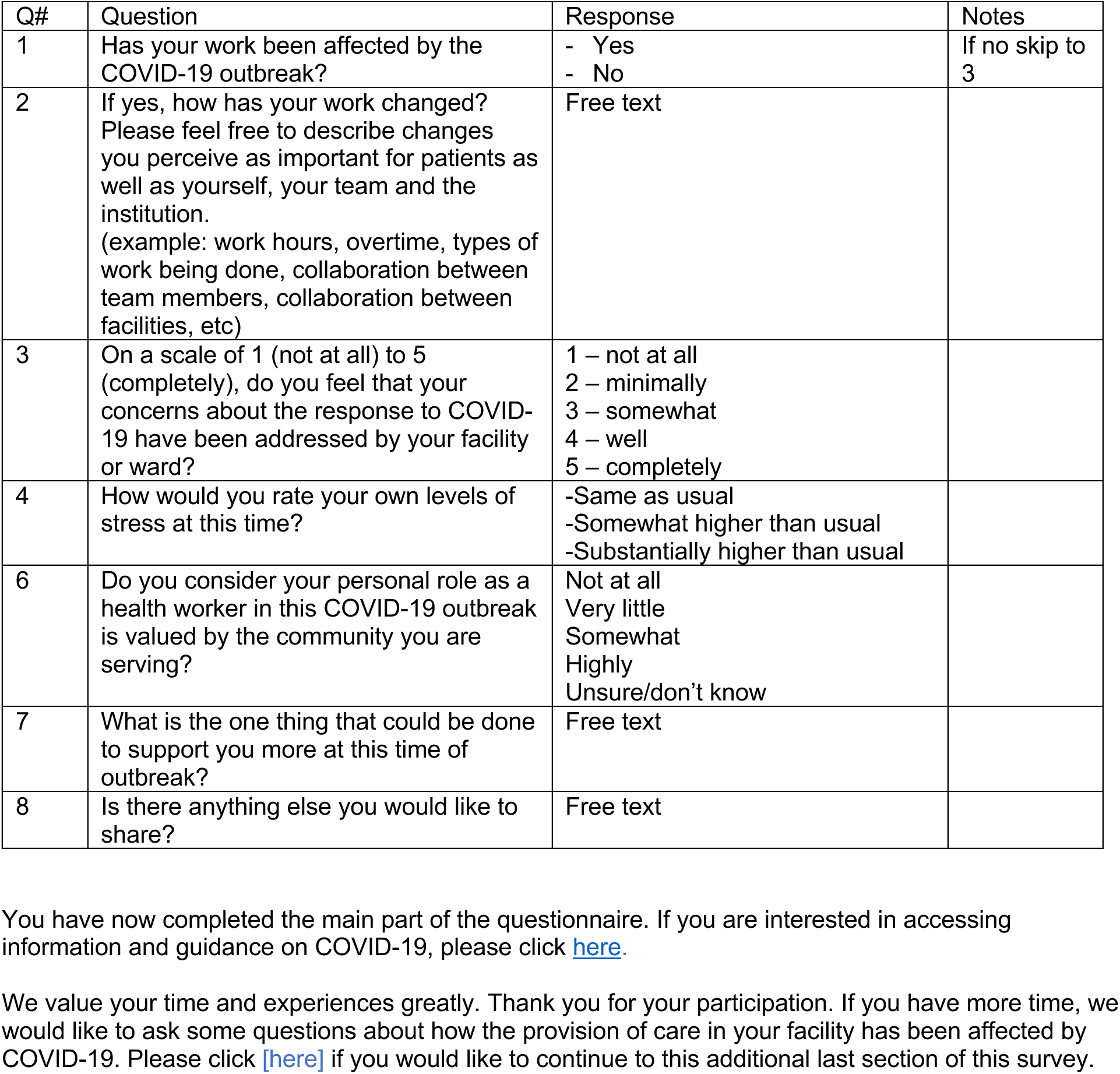
Your work and experience in light of the COVID-19 outbreak.

Additional module. Effect of COVID-19 on the provision of maternal and newborn care. Can you describe how the COVID-19 outbreak has affected the provision of care to women and newborns in your facility and community? This includes changes made directly in response to the threat of COVID-19 and other indirect influences (for example, pressure on the health system).

| Q# | Question | Response |
| --- | --- | --- |
| 1 | Changes to provision of **outpatient antenatal** care (examples include staffing levels, location – in person or phone/internet, waiting times, patient flow, ability to conduct all routine tests and investigations, etc) | Yes - No<br>If yes, please describe the changes |
| 2 | Changes to provision of **inpatient antenatal** care | Yes - No<br>If yes, please describe the changes |
| 3 | Changes to capacity to provide **intrapartum care** (number of rooms or beds, availability of equipment, supplies and medications) | Yes - No<br>If yes, please describe the changes |
| 4 | Changes to rules on number or type of **labour companions** (includes family members and professional doulas, whether able to stay overnight, etc) | Yes - No<br>If yes, please describe the changes |
| 5 | Changes to **pain relief** options available to women in labour | Yes - No<br>If yes, please describe the changes |
| 6 | Changes to **rules on induction** of labour | Yes - No<br>If yes, please describe the changes |
| 7 | Changes to **caesarean section provision** (e.g. location of theatre, type of anaesthesia, guidelines for elective csections) | Yes - No<br>If yes, please describe the changes |
| 8 | Changes to **visiting hours** or number/type of visitors, for mothers and newborns | Yes - No<br>If yes, please describe the changes |
| 9 | Changes to **inpatient postnatal** care provision following vaginal births (for example, shorter length of stay, frequency of routine checks) | Yes - No<br>If yes, please describe the changes |
| 10 | Changes to **inpatient postnatal** care provision following caesarean section births (for example, change in cleaning schedules, hand hygiene indications, hand hygiene supplies, delivery equipment decontamination and sterilisation) | Yes - No<br>If yes, please describe the changes |
| 11 | Changes to **outpatient postnatal** care provision (ability to follow-up in women's homes, breastfeeding counselling, postpartum family planning etc) | Yes - No<br>If yes, please describe the changes |
| 12 | Changes to provision of **routine newborn** care before discharge (screenings, vaccinations, etc) | Yes - No |
|  |  | If yes, please describe the changes |
| 13 | Changes to provision of **newborn intensive** care (example: bed capacity, oxygen, equipment) | Yes - No<br>If yes, please describe the changes |
| 14 | Changes to provision of **non-essential care** (such as cancellations of elective surgery, gynecological procedures, IVF provision, etc) | Yes – No<br>If yes, please describe the changes |
| 15 | Changes to **staffing levels or team/shift composition** (for example, more people hired, such as cleaners, change in shift hours, staff being absent due to symptoms ) | Yes - No<br>If yes, please describe the changes |
| 16 | Changes to process of **referring patients to other facilities** (timing, destination, mode of transport, capacity, guidelines) | Yes - No<br>If yes, please describe the changes |
| 17 | Changes to **process of receiving incoming maternity referrals**, including antenatal and emergency. (timing, destination, mode of transport, capacity, guidelines) | Yes - No<br>If yes, please describe the changes |
| 18 | Do you feel that **women's use of care** in your facility has been affected by COVID-19? For example, do you see fewer or more patients than usual? If so, why? | Yes - No<br>If yes, please describe the changes |

**Supplementary File 2.**
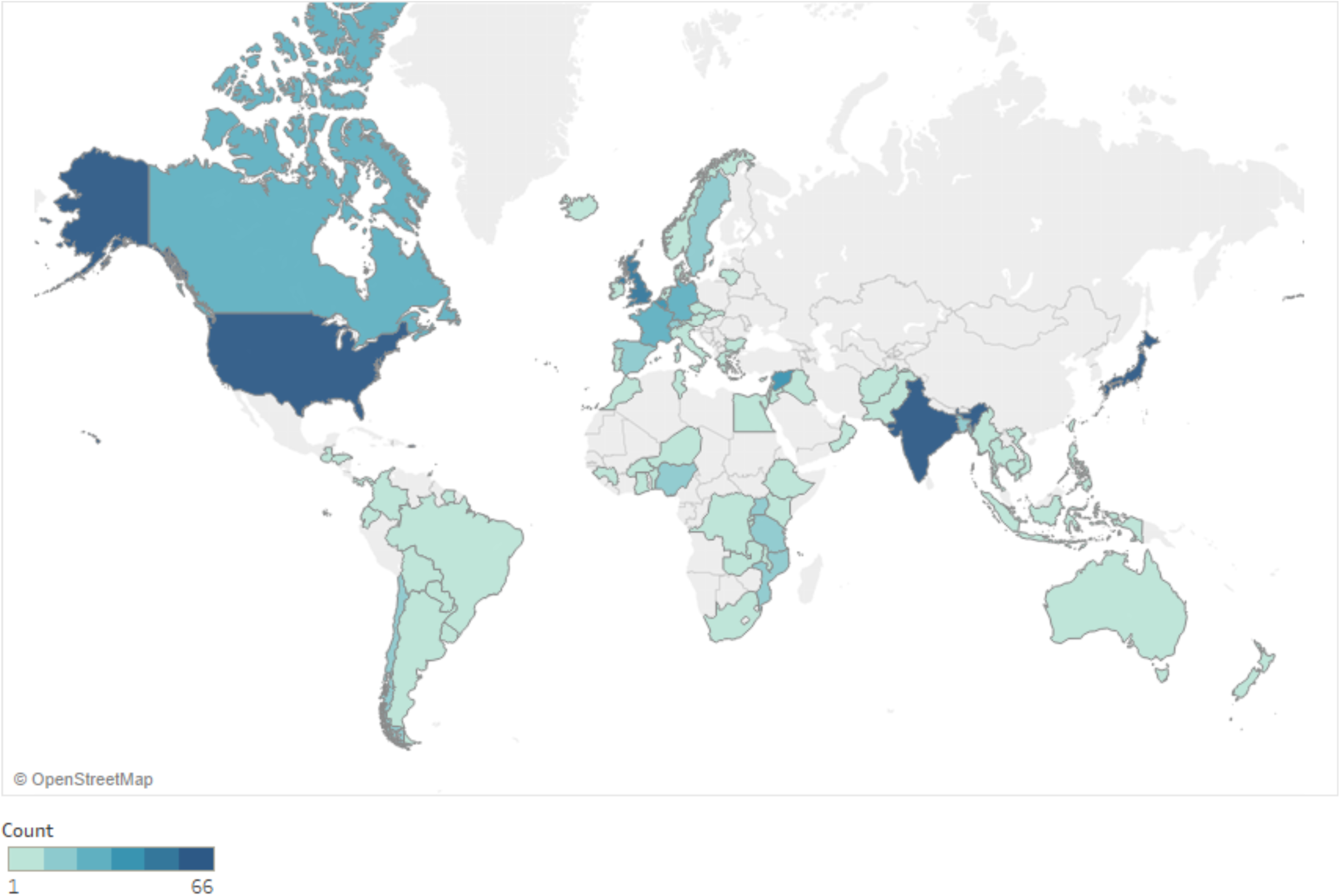
Frequency distribution of respondents by country

